# Assessing Age-Specific Vaccination Strategies and Post-Vaccination Reopening Policies for COVID-19 Control Using SEIR Modeling Approach

**DOI:** 10.1101/2021.02.18.21251981

**Authors:** Xia Wang, Hulin Wu, Sanyi Tang

**Affiliations:** School of Mathematics and Information Science, Shaanxi Normal University, Xi’an, People’s Republic of China; Department of Biostatistics and Data Science, University of Texas Health Science Center at Houston, Houston, USA

**Keywords:** age-structure, vaccination strategy, COVID-19 control, reopening policy, social contact

## Abstract

**Background:** As the availability of COVID-19 vaccines, it is badly needed to develop vaccination guidelines to prioritize the vaccination delivery in order to effectively stop COVID-19 epidemic and minimize the loss.

**Methods:** We evaluated the effect of age-specific vaccination strategies on the number of infections and deaths using an SEIR model, considering the age structure and social contact patterns for different age groups for each of different countries.

**Results:** In general, the vaccination priority should be given to those younger people who are active in social contacts to minimize the number of infections; while the vaccination priority should be given to the elderly to minimize the number of deaths. But this principle may not always apply when the interaction of age structure and age-specific social contact patterns is complicated. Partially reopening schools, workplaces or households, the vaccination priority may need to be adjusted accordingly.

**Conclusions:** Prematurely reopening social contacts could initiate a new outbreak or even a new pandemic out of control if the vaccination rate and the detection rate are not high enough. Our result suggests that it requires at least nine months of vaccination before fully reopening social contacts in order to avoid a new pandemic.

## Introduction

In 2020, novel coronavirus (SARS-CoV-2) pneumonia broke out in the world. As of December 28, 2020, a total of 80.724 million confirmed cases and 1.764 million deaths have been reported worldwide, according to the data from Johns Hopkins University website[1]. The cases infected by the new variants of the SARS-CoV-2 have been confirmed in many countries, including the UK, South Africa, France, Japan, Thailand, Canada, Portugal and so on. Great efforts in the world have been made for controlling COVID-19 pandemic in the past year, but unfortunately, the COVID-19 epidemic is still deteriorating in many countries.

In such a severe situation, the development and use of vaccines have been a great hope to control COVID-19 epidemic. There are over 100 COVID-19 vaccines under development in the past year. It is gratifying that a great progress has been made in vaccine development, some vaccines have been approved to use and some others are undergoing phase 3 clinical trials[2]. Many countries are preparing to implement and deliver vaccination sequentially based on some priority guidelines. In the early stage of COVID-19 epidemic, the main research focus is to assess the basic reproductive number, infection scale and the impact of population mobility on COVID-19 transmission [3-6]. With the wide spread of the epidemic, the research focus was quickly shifted to develop and evaluate the effectiveness of control measures[7-11]. The development of COVID-19 vaccinesis accelerated, but the capacity of vaccine production is limited and it may take time to make vaccine available to all the people who are willing to receive it. Thus, it is badly needed to develop vaccination strategies in order to maximize the benefit of vaccination in controlling COVID-19 epidemic.

Recently in several modelling studies[12-16], researchers have tried to investigate the optimal control of vaccination in mitigating the epidemics. Tang et al. investigated the reopening strategies for different countries assuming the availability of vaccines[15], and they suggested that country level-based reopening strategies should be considered, according to the quarantine rate, testing ability, and the condition of vaccines. Considering the limitation of initial supply of SARS-CoV-2 vaccines, five vaccine prioritization strategies are examined for the US using an age-stratified SEIR model[14]. In fact, the age has been shown to be an important factor for susceptibility and death rate[17-20], thus, the age structure plays an important role in vaccination strategies. We noticed that different countries have different age structures and different contact networks among different age groups [21]. Based on this observation, we extended the age-structured SEIR model to identify the optimal age-specific vaccination distributions for different countries with different age structures and different contact networks. In addition, we further explored the post-vaccination reopening policies based on the model with the optimal vaccination distributions.

## Methods

### Model

We propose the following age-structured SEIR model:

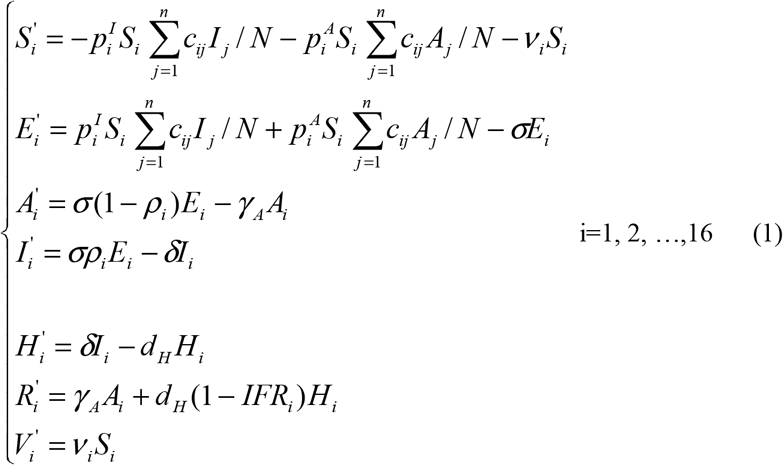

In order to consider the effect of different age-specific vaccination strategies to control the epidemic in different countries (e.g., China, India and Italy), we divided the whole population into 16 age groups(0-4,5-9,10-14,…,75+), according to the contact data[21]. We also divided the population as the susceptible(S), the exposed(E), the asymptomatic infected(A), the symptomatic infected(I), the hospitalized(H), the recovered(R) and the vaccine (V). We assumed that people with different ages had different susceptibility 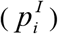, and the proportion of asymptomatic infections (*ρ*_*i*_) also depended on ages[22]. We also assume that the asymptomatic infected individuals are less infectious compared to the symptomatic infected individuals 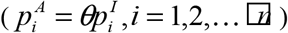 σ is the transition rate from the exposed to the infected, *γ* _*A*_ and *d*_*H*_ (1− *IFR*_*i*_) are recovery rates, *d*_*H*_*IFR*_*i*_ is the death rate of age group *i, IFR*_*i*_ is the age-specific infection fatality rate for age group *i* [25], *v* _*i*_ is effective vaccination rate of age group *i*, and *c*_*ij*_ is the contact rate of age group *j* by age group *i* [21]. *c*_*ij*_ and *δ* are assumed to be time varying functions (see more details in the Appendix).

In order to compare the impact of different vaccination strategies on the epidemic, we considered two vaccination strategies: one is the uniform vaccination strategy, and the other is the age-specific vaccination strategy. In the case of the uniform vaccination strategy, we assume *v*_1_ = *v* _2_ = … = *v*_16_ = *v*. For the age-specific vaccination strategy, we assume that *v* _*i*_ = *Κp*_*i*_, *i* = 1,…,16, where *Κ* is a scaling factor and *p*_*i*_ is vaccination age distribution. In order to maintain the consistency of the number of vaccinations everyday, We assume 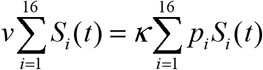. Considering that Beta distribution is defined in a finite interval and its density function is very flexible (it can be either unimodal or U-shaped).Moreover, the uniform distribution is a special case of Beta distribution. So, to reduce the number of parameters, we assume that the vaccination age distribution follows a Beta distribution, *Beta*(*α, β*) with parameter *α* and *β*, i.e., *p*_*i*_ = *F* (*i* /16 | *α, β*) − *F* ((*i* − 1) /16 | *α, β*), where *F* (*x* | *α, β*) is the Beta cumulative distribution function.

The basic reproduction number can be calculated according to [23], which is the principal eigenvalue of the following matrix Λ (see more details in the Appendix):

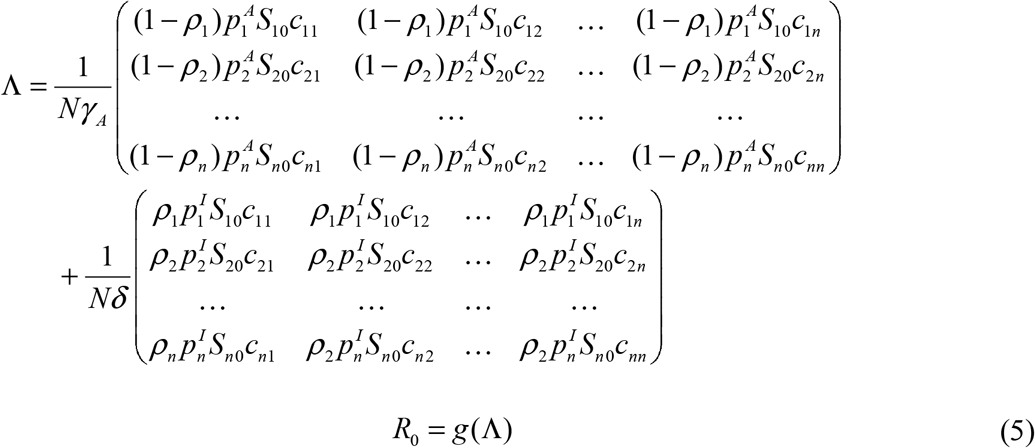

where g(.) denotes the spectral radius of a matrix.

### Data and parameter calibration

Three countries (China, Italy and India) with different population age distributions(Fig. 2) are chosen to be analyzed in this study. The data for the population age distribution are obtained from https://www.populationpyramid.net/japan/2019/ (Fig.S1) [24]. The data of daily reported COVID-19 cases were obtained from https://github.com/CSSEGISandData. We parameterize the contact matrices at the initial time for each country using the age-dependent contact rates(households 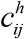. workplaces 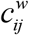, schools 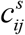 and other locations 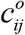) estimated by Prem et al.[21]. The summation of the contacts across different sites was used to be the baseline contact matrix 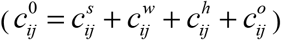 (Fig.S2). The susceptibility of different age groups 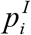 can be derived from Keeling et al. [20]. According to Prem et al.[22], we also assume that the younger individuals are more likely to be asymptomatic, so the proportion of asymptomatic infections (*ρ*_*i*_) is assumed to be 0.4 when *i* ≤ 4, and 0.8 when *i* > 4 [22]. Moreover, we assume that the infectiousness of asymptomatic is less than that of the symptomatic individuals, and *θ* is assumed to be 0.25[22]. The parameter values related to COVID-19 and its transmission are derived from related references, and other parameters are estimated by fitting model to data(Table 1 and Appendix).

**Table 1:** Parameter definitions and estimation for the age structured SEIRmodel.

| Parameter | Definitions | China | Italy | India | Sources |
| --- | --- | --- | --- | --- | --- |
| $c_{ij}^0$ | Contact rate of age class $i$ with age class $j$ at the initial time | - | - | - | Prem et al.,2017 |
| $c_{ij}^b$ | Minimum contact rate of age class $i$ with age class $j$ | - | - | - | Estimated |
| | | | $0.0212(r_{c_1})$ | | |
| $r_c$ | an exponential decrease rate | 0.1216 | $0.0202(r_{c_2})$ | 0.01 | Estimated |
| | | | $0.0571(r_{c_3})$ | | |
| | | | $0.3096(q_{c_1})$ | | |
| $q_c$ | The proportion between $c_{ij}^b$ and $c_{ij}^0$ | 0.1 | $3(q_{c_2})$ | 0.3342 | Estimated |
| | | | $0.3001(q_{c_3})$ | | |
| $t_c$ | Switching time of control strength | 16 | 0 | 0 | Assumed |
| $t_{c_1}(t_{c_2})$ | Switching time of control strength | - | 153(268) | - | Estimated |
| $p_i^I$ | Probability of infected individuals transmission per contact | - | - | - | Keeling2020 |
| $p^A$ | Probability of asymptomaticinfected individuals transmission per contact | - | - | - | |
| $\theta$ | Scaling factor | 0.25 | 0.25 | 0.25 | Prem et al., 2020 |
| $\nu$ | Vaccination rate | | | | Varied according to scenario |
| $\rho_i$ | Proportion of symptomatic infection | 0.4 for $i \leq 4$, 0.8 for $i > 4$ | 0.4 for $i \leq 4$, 0.8 for $i > 4$ | 0.4 for $i \leq 4$, 0.8 for $i > 4$ | Prem et al., 2020 |
| $\sigma$ | Transition rate of exposed individuals to the infected class | 1/5 | 1/5 | 1/5 | Tang2020 |
| $\delta_0$ | Diagnosis rate of infected individuals at the initial time | 0.07 | 0.1267 | 0.1573 | Estimated |
| $\delta_f$ | Maximum diagnosis rate of infected individuals | 0.4 | 0.2974 | 0.2522 | Estimated |
| $r_\delta$ | an exponential increase rate | 0.0518 | 0.0403 | 0.3 | Estimated |
| $q_\delta$ | The proportion between $1/\delta_f$ and $1/\delta_0$ | 5.7143 | 2.3480 | 1.6029 | Estimated |
| $\gamma_A$ | Recovery rate of asymptomatic infected individuals | 0.18 | 0.18 | 0.18 | Wang et al., 2020 |
| $d_H$ | Removal rate of hospitalized/confirmed infected individuals | 0.2 | 0.2 | 0.2 | Bubar et al., 2020 |
| $IFR_i$ | Infection fatality rate of age group $i$ | - | - | - | Levinet al., 2020 |
| $S_{i0}$ | Initial susceptible population of age group $i$ | - | - | - | Data |
| $E_{i0}$ | Initial exposed population of age group $i$ | 10 | 1 | 1 | Estimated |
| $I_{i0}$ | Initial symptomatic infected population of age group $i$ | 37.2457 | 60.8411 | 1 | Estimated |
| $A_{i0}$ | Initial asymptomatic infected population of age group $i$ | 30 | 1 | 1 | Estimated |

**Fig 1:**
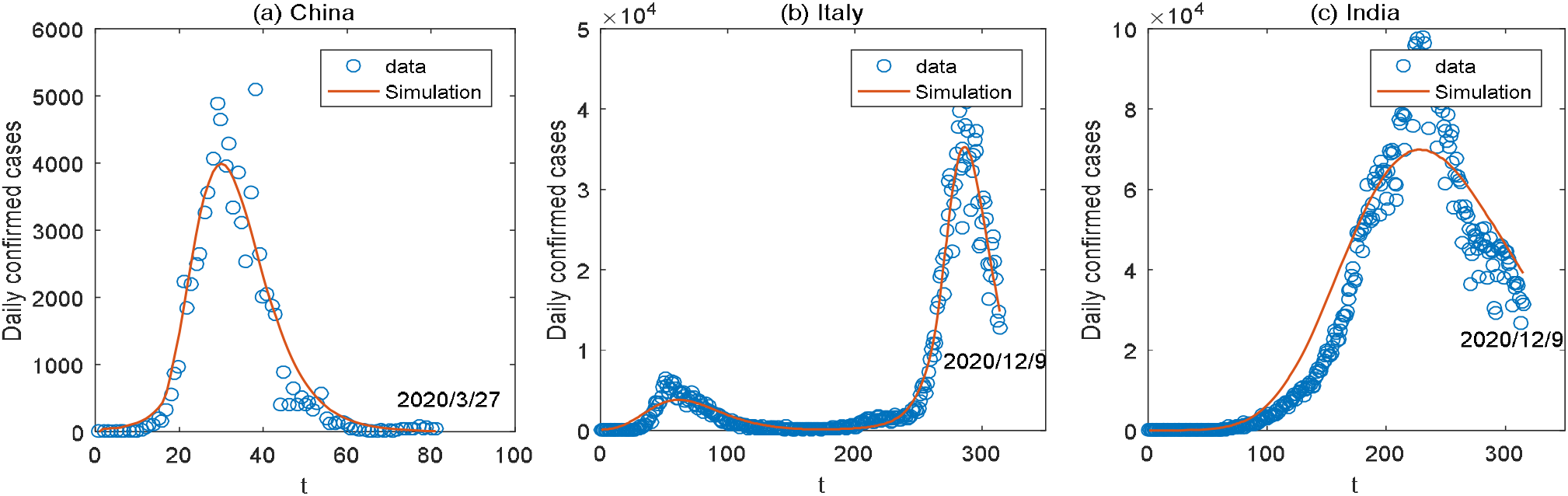
Observed daily new cases(dots) and model fitting results (solid curve) for China (a), Italy (b) and India (c).

**Fig 2:**
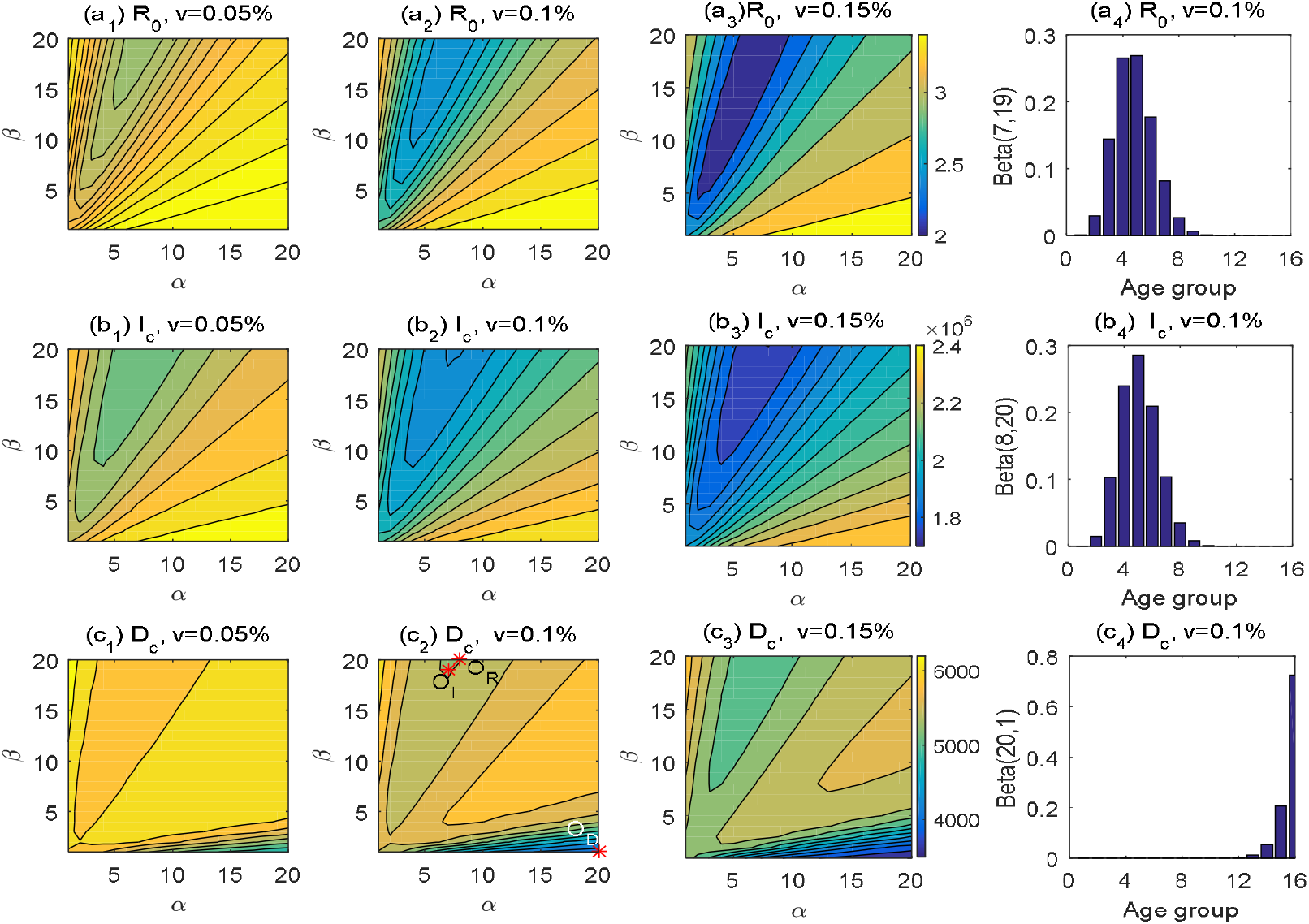
The contour plot of the three endpoints: the basic reproduction number (R_0_, 1^st^ row), the cumulative number of infections (I_c_, 2^nd^ row) and the cumulative number of deaths (D_c_, 3^rd^ row) for India. The optimal age-specific vaccination distributions for these three endpoints are shown in (a_4_), (b_4_) and (c_4_)respectively when *v* = 0.1%. *O*_*R*_, *O*_*I*_,and *O*_*D*_ are the optimal points obtained by minimizing the three endpoints (R_0_, I_c_, D_c_)respectively.

### Evaluation of the optimal age-specific vaccination distribution

The basic reproduction number (R_0_), the cumulative number of infections (I_c_) and the cumulative number of deaths (D_c_) are the key indicators of the severity of infectious diseases and public health concerns, and thus are used as the endpoints or outcomes to evaluate the effectiveness of different vaccination strategies in this study. Since the Beta distribution of the age-specific vaccination can be uniquely determined by the parameters *α* and *β*, we will evaluate each of the three endpoints as a function of the two parameters. We assume that the vaccine will be delivered continuously for 180 days in a fixed rate. The optimal *α* and *β*, i.e., the optimal age-specific vaccination distribution(OAVD), can be determined by minimizing each of the three endpoints from the vaccination initiating time (T) to T+180 days for each country. We used interior-point method to optimize the three endpoint functions. Considering that the epidemic in China has been well controlled by now and the sporadic outbreaks in different cities are mainly due to overseas imports, we assume that all people in China are susceptible and one infected individual is imported from overseas to potentially initiate a new epidemic. The detection rate and the contact rate are fixed to be *δ*_*f*_ and 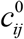 in our model evaluations.

## Results

### The optimal age-specific vaccination distribution

We evaluated the effect of different age-specific vaccination distributions on the control of COVID-19 epidemic, so that we could determine the OAVD by minimizing the aforementioned three endpoints for different countries (China, India and Italy)respectively. For each case, we set the vaccination rate *v* as 0.05%, 0.1% and 0.15%respectively. Fig.2 shows the OAVDs and the contour plot of the three endpoints as a function of *α* and *β* for India. We can see that the OAVD obtained by minimizing R_0_ and I_c_ are similar when *v* = 0.1% (Fig.2(*a*_4_)and(*b*_4_)). This suggests that for India, the priority of vaccination should be given to teenagers and young people, i.e., those around 10-34 years old, in order to minimize R_0_ and I_c_. This is presumably because R_0_ is directly related to the spread of infections in the SEIR model. Thus, the OAVD for these two endpoints is also similar for other vaccination rate (Table 2 and Table S1). However, the effect of OAVDs on D_c_ can be significantly different from that for other two endpoints (Fig. 2). In order to minimize D_c_, the OAVD (Fig. 2(c_4_)) suggests that the high priority should be given to elders in India. Notice that the effect of age vaccination distribution on D_c_ is complicated and there are possible multiple solutions for the OAVD. For example, when the vaccination rate is 0.15%, the contour plot for D_c_ (Fig.2 (*c*_3_)) shows two troughs with two optimal age-specific vaccination distributions, and both solutions could keep D_c_ under 5000. One solution (Beta(7,20)) is similar to that for R_0_ and I_c_, and the other is when *β* is very small and *α* is very large, i.e., (Beta(20,1)). In general, the vaccination priority should be given to the elderly in order to control D_c_, since the death rate is much higher for elderly[25]. However, giving priority to young people can also reduce D_c_ to less than 5000, while also keeping the total infection number slow in India. We also noticed that it is not effective to control COVID-19 epidemic to give vaccination priority to children for all the cases that we have considered. We also observed that the effect of age-specific vaccination distribution on the three endpoints is larger when the vaccination rate is higher (Fig. 2, Fig. S3 and Fig. S4). For the case in India, the optimal age-specific vaccination strategy can reduce I_c_ by 177,270(8.5%) or reduce D_c_ by 1,514 (27%) compared to that of the uniform vaccination strategy.

**Table 2:**
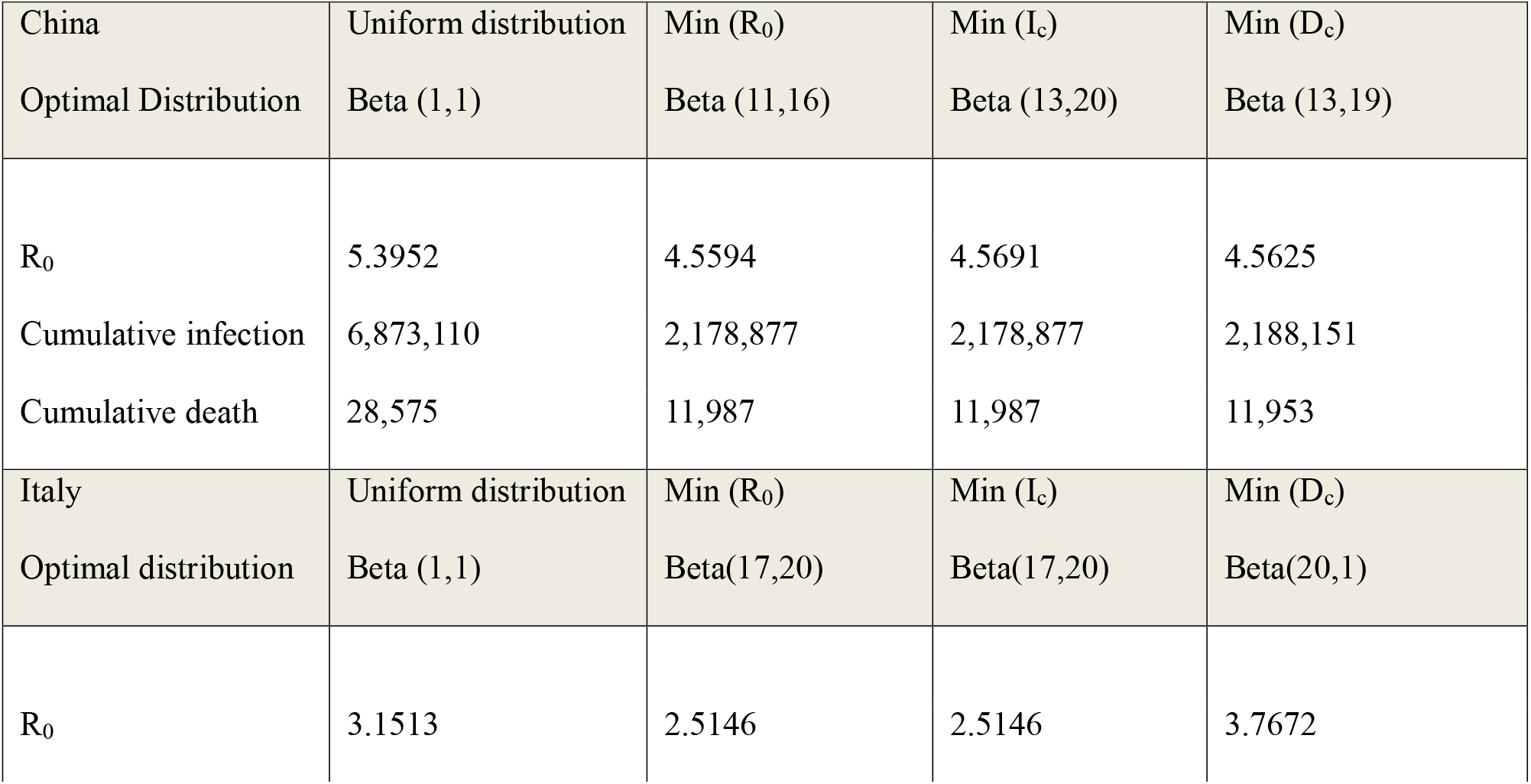
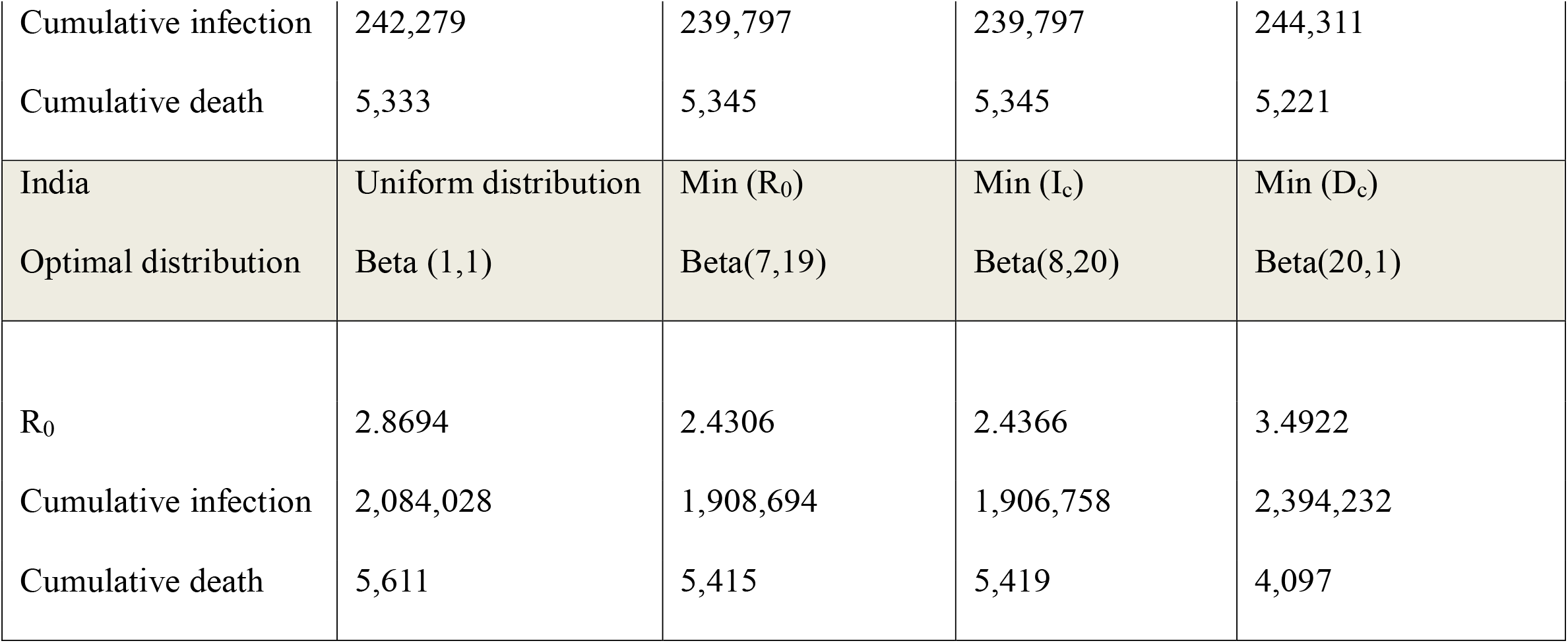
The final outcomes with the optimal age-specific distributions vs.the uniform distribution (*v* = 0.1%)for the three countries (China, Italy and India)

| China | Uniform distribution | Min ($R_0$) | Min ($I_c$) | Min ($D_c$) |
| --- | --- | --- | --- | --- |
| Optimal Distribution | Beta (1,1) | Beta (11,16) | Beta (13,20) | Beta (13,19) |
| $R_0$ | 5.3952 | 4.5594 | 4.5691 | 4.5625 |
| Cumulative infection | 6,873,110 | 2,178,877 | 2,178,877 | 2,188,151 |
| Cumulative death | 28,575 | 11,987 | 11,987 | 11,953 |
| Italy | Uniform distribution | Min ($R_0$) | Min ($I_c$) | Min ($D_c$) |
| Optimal distribution | Beta (1,1) | Beta(17,20) | Beta(17,20) | Beta(20,1) |
| $R_0$ | 3.1513 | 2.5146 | 2.5146 | 3.7672 |
| Cumulative infection | 242,279 | 239,797 | 239,797 | 244,311 |
| Cumulative death | 5,333 | 5,345 | 5,345 | 5,221 |
| India | Uniform distribution | Min ($R_0$) | Min ($I_c$) | Min ($D_c$) |
| Optimal distribution | Beta (1,1) | Beta(7,19) | Beta(8,20) | Beta(20,1) |
| $R_0$ | 2.8694 | 2.4306 | 2.4366 | 3.4922 |
| Cumulative infection | 2,084,028 | 1,908,694 | 1,906,758 | 2,394,232 |
| Cumulative death | 5,611 | 5,415 | 5,419 | 4,097 |

The effect of age vaccination distributions on the three endpoints in Italy is similar to that in India (Fig. S2), which resulted in similar OAVDs. Thus, the vaccination priority should be given to young and middle aged people in order to control I_c_ while the priority should be given to the elderly in order to control D_c_. However, for the case of China, the effect of age vaccination distributions on R_0_ and I_c_ is similar to that in India and Italy (Fig. S4), which resulted in similar conclusions to control I_c_; but the effect of age vaccination distributions on D_c_ is different from other two countries. The OAVD obtained by minimizing D_c_ is also similar to that of other two endpoints (see Fig. S4(c_4_)). Interesting, we also observed that the OAVD has only a small effect on I_c_ and D_c_ in Italy when *v* = 0.1% (Table 2). However, the optimal age-specific vaccination strategy has a big effect on I_c_ and D_c_ for the case of China. Use of the optimal age-specific vaccination strategy could reduce I_c_ from 6,873,110 to 2,178,877 or more than 3-fold reduction or it can reduce D_c_ from 28,575 to 11,953 (2.4-fold reduction) compared to that of uniform distribution in China (Table 2).

### Optimal reopening strategies to restore social contacts with vaccination

After COVID-19 vaccines are administered among the population, it is expected to reopen or even gradually restore social contacts as normal as that before the COVID-19 epidemic. Here we further explore the effect of gradual reopening polices from contact control measures under the optimal age-specific vaccination strategy obtained by minimizing I_c_. We simulated the total number of infections during a time period of 6 months under different degrees of contact control release and for different daily vaccination rates =0∼0.15%with the initiation time of vaccination as *T* + 1. We considered the following scenarios of contact control release at different sites:

a. Reopen the schools: 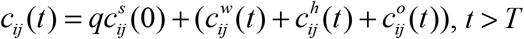
b. Reopen the workplace: 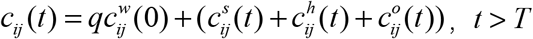
c. Reopen home: 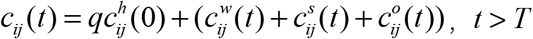
d. Reopen others: 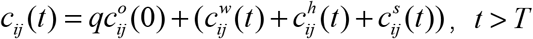
e. Reopen all above: 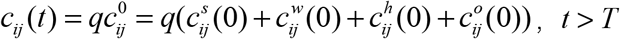

where *q*_*c*_ < *q* ≤ 1. Notice that the contact control is completely released (restored to normal life) when *q* = 1

Fig.3 shows the simulation results for I_c_ (in log10 scale) for India during the time period of 180 days [T+1,T+180] with different vaccination rates*v* (ranging 0-1.5%) and different degrees of reopening *q* (ranging 0 to 1) under different releasing times, T+1, T+30 and T+60. From Fig.3, we can see that *q* has a significant effect on the number of infections. For the case of releasing all contact controls completely, I_c_ may exceed 100 million in 6 months in India if *q* > 0.7 without vaccination, i.e., *v* = 0 (Fig.3(e1)). Moreover, even with vaccination initiated, it may result in a high I_c_ if the contact control measure is released too early, i.e., early release for one month may lead to almost 10-fold difference in the number of infected people in India (see Fig.3(e2) and Fig.3(e3)). For the cases of partial contact control release at different sites, releasing “other contacts” brings the highest number of infected people, followed by releasing schools and households. The risk of releasing contacts in workplace is minimal.

**Fig 3:**
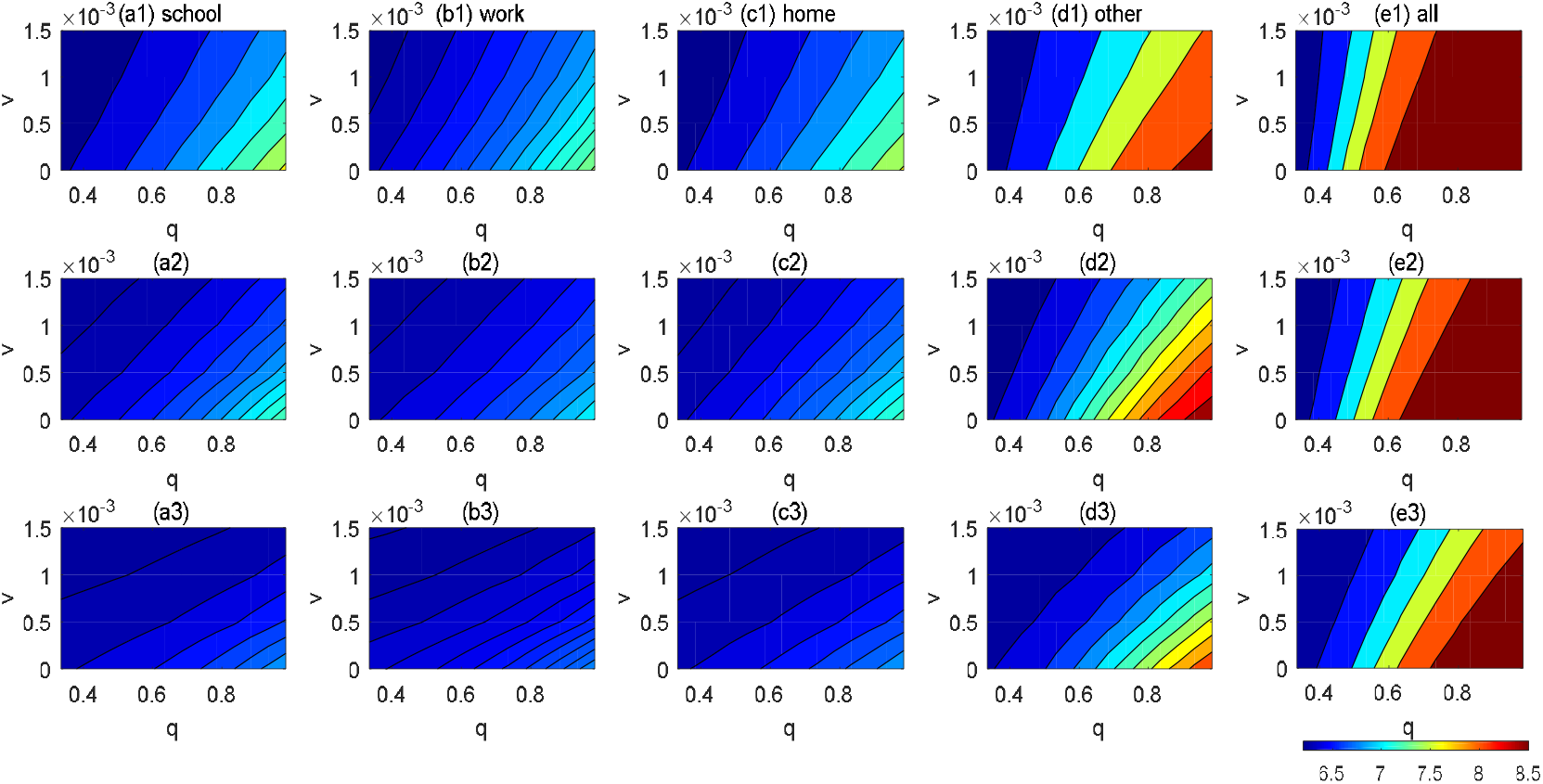
Simulation results for the number of infections (in log10 scale) for India during the time period [T+1,T+180]for five different scenarios of contact control release (a-e)for different values of *q* and *v* under different releasing times,T+1 (1^st^row), T+30(2^nd^row), and T+60 (3^rd^row).The optimal age-specific vaccination strategy with the initiation time of vaccination as *T* + 1 was assumed for all the simulation scenarios.

For the case of Italy (Fig. S5), the effect of different releasing strategies and releasing times is similar to that in India. In addition, reopening schools would have a small effect on I_c_ compared to that reopening other sites(Fig.S5). For the case of China, we investigated I_c_ after reopening different sites if one infected case would be imported(shown in Fig.S6).When the detection rate is high, δ=0.4, partially reopening some sites will not cause a pandemic, but thousands of people could be infected (Fig.S6) if the reopening rate is high and vaccination rate is low, say, *q* > 0.9 and *v* ≤ 0.05%. However, if the detection rate is low, δ=0.1, it might cause hundreds of thousands of people to be infected within 6 months if all the sites would be reopened.

Considering that partially reopening different sites or locations for social contacts may lead to changes of the OAVD, we evaluated the effect of partially reopening different sites on the OAVDs. In Fig. 4, we show the OAVDs for the endpoint of R_0_ under different partially reopening policies. We assumed the vaccination rate ν=0.1%. From Fig.4, we can see that the vaccination priority should be given toward younger people (5-20 years old) in China if the social contacts are just open for schools or households, while the vaccination priority might need to be given to older people (around 40 years old) in China if the workplace is open first (Fig. 4(a)). For the case of Italy, the vaccination priority might need to shift toward younger population if the household is opened, but reopening schools and workplaces in Italy do not have much effect on the optimal age distribution (Fig. 4(b)). For the case of India, the vaccination priority might need to shift toward older population if the workplace is opened, but reopening schools and households in India has little effect on the OAVD (Fig. 4(c)). The differences in the effect of partially reopening different sites on the OAVDs for different countries are presumably due to the differences in age structures of their population (Fig. S1) and social contact patterns (Fig. S2).

**Fig 4:**
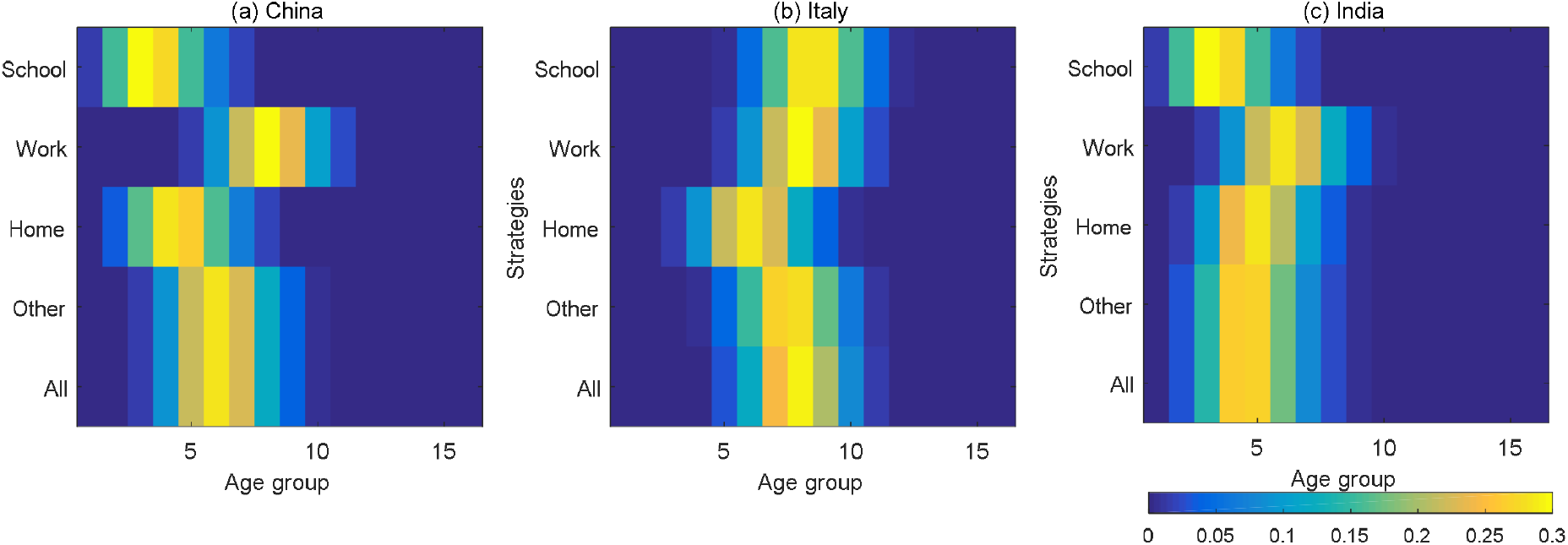
Heat map of the optimal age-specific distribution under different reopening strategies. The vertical coordinates represent different reopening strategies: only release contacts in schools, only release contacts in workplaces, only release contacts in households, only release contacts in other locations and reopen all.

If the post-vaccination reopening is too fast and the detection rate is not high enough, it is possible to cause a new outbreak of COVID-19. Based on our model, we examined R_0_ as a function of the reopening rate (*q*) and detection rate (δ) under the OAVD. The contour plots of R_0_ for the three countries (India, Italy and China) are shown in Fig.5. Fig.S7 and Fig.S8 respectively. From Fig.5 for the case of India, the estimated value of δ=0.2522, q=0.3342 and R_0_<1, thus, the epidemic is under control. However, if δ is kept as the same and the vaccination lasts for 3 months with the vaccination rate ν=0.1%, q is increased to >0.5, it would result in R_0_>1, which would cause a new pandemic (Fig.5(a)). However, if the vaccination lasts longer with a higher vaccination rate, the epidemic is still under check. For example, if the vaccination could last for 12 months with ν=0.15%, the full reopening(q=1) could not cause a new outbreak(Fig.5(d)) in India (R_0_<1) if δ is kept as the same. For the case of Italy, a similar trend is observed, but it looks safe to fully open the social contacts if the vaccination could last for 6 months or longer with a higher vaccination rate ν>0.1%, since its detection rate is higher (Fig. S7). For the case of China, it is similar to Italy, it also requires 6 months or longer vaccination period with a vaccination rate ν>0.1% to be safe for fully reopening social contacts (Fig. S8).

**Fig 5:**
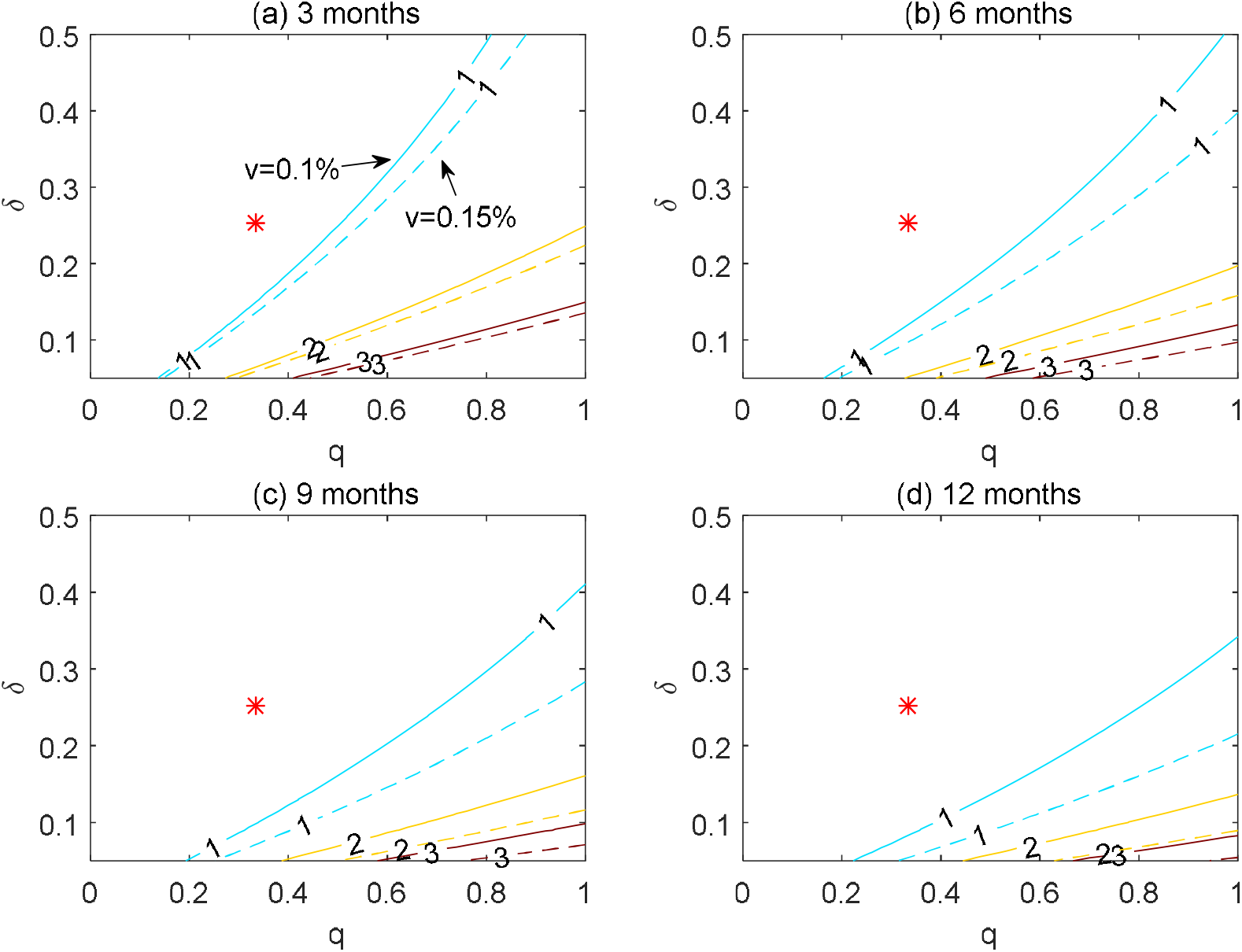
Contour plot of the basic reproduction number (R_0_) as a function of the contact control release rate (q) and the detection rate (δ) with different daily vaccination rates v=0.1% (the solid line) and v=0.15%(the dash line) under the optimal age-specific vaccination strategy in India. The vaccination periods are set to be 3 months(a), 6 months(b), 9 months (c) and 12 months (d). The red dot is the current value of these two parameters.

### Conclusions and Discussions

The COVID-19 is still widely spreading around the world and many people are killed every day. Vaccines offer a great hope for end to COVID-19 pandemic, but an effective vaccination strategy is badly needed in order to quickly stop the epidemic and restore the normal life of people. Due to the limited manufacturing capacity, COVID-19 vaccines may not be immediately available to all the people who are willing to receive. It is necessary to develop priority guidelines for different groups of people to receive vaccines sequentially. Intuitively the high risk populations such as first responders, elderly and people with high-risk health conditions should receive the vaccines first[26].The question is, how to prioritize the rest of the population for vaccination after the high risk population. In this study, we used the SEIR modeling approach to investigate the optimal age-specific vaccination strategies. In particular, we also utilized the information of the age structure of the population (Fig.S1) and contact network (Fig.S2) data in order to optimize the age-specific vaccination strategy for different countries. Compared with the previous literature[14,16], we used a continuous function, Beta distribution, to approximate the age-specific vaccination distribution, so that we could conveniently optimize the age distribution by minimizing the three endpoints with respect to the Beta distribution parameters(α and *β*).

Our results show that the OAVD and the vaccination priority are different for different outcomes. To minimize D_c_, the vaccination priority should be given to the elderly, while the vaccination priority should be given to socially active younger people if the goal is to minimize I_c_. This general principle can be different for different countries since the age structure of the population and social contact patterns are different for different countries (Fig.S1 and Fig.S2). For example, the vaccination priority given to the middle-age people could minimize both I_c_ and D_c_ in China, presumably due to the complicated interactions between the age structure of the population and the social contact patterns. The results based on R_0_ and I_c_ are similar, presumably because R_0_ is directly related to the spread of infections.

With availability of effective vaccines, we expect to quickly restore normal life with regular social contacts. However, prematurely reopening social contacts during the vaccination stage may cause large I_c_ and D_c_, even initiate a new pandemic. A good strategy is to gradually or partially reopen some necessary sites or locations with a phased plan. Our simulation results show that it is safer to partially reopen the schools and workplaces, instead of households and other locations for the case in India (see Fig.3). In addition, the optimal age-specific vaccination strategy needs to be adjusted accordingly if this is the case. If the schools are reopened, the vaccination priority should be shifted more toward children and teenagers; if the workplaces are reopened, the vaccination priority should be shifted more toward middle-aged people in India (see Fig.4). It seems that, if all the sites are fully reopened for social contacts too early, it may cause a large number of infections (Fig.3) or even initiate a new pandemic, possibly out of control (R_0_>1) in India if the vaccination rate and detection rate are not high enough (Fig.5). For the cases of Italy and China, similar general principles can be applied, but the detailed reopening strategies need to be tailored due to the differences in age structures of the population and social contact patterns.

In conclusion, our results show that the age structures of the population and social contact patterns have a significant impact on the effect of age-specific vaccination strategies. In the case of limited vaccine resources, it needs to consider different age-specific priority guidelines for general population in order to control the COVID-19 pandemic more effectively. Moreover, different countries need to develop specific vaccination strategies according to the age structures of their population and social contact patterns. Our established age-structured SEIR models can also be used to evaluate post-vaccination reopening policies in order to safely restore to normal life. To obtain the OAVD, we minimized the three endpoints. Although it may fall on the boundary of the search space of parameters (α,β) (Fig.2 and Table 2), sensitivity analysis results show that it won’t affect the priority of vaccination population. The proposed optimal age-specific vaccination strategies can be implemented in the vaccination scheduling system, that requires additional programming effort, compared to the simple uniform distribution or first-come-first-served strategies. But it worths the effort if it can save thousands of lives or reduce the large number of infections.

## Data Availability

All data refferred to in the manuscript is shown in the manuscript.

https://www.populationpyramid.net/japan/2019/

https://github.com/CSSEGISandData

## Notes

## Acknowledgments

We thank the editors and reviewers.

## Author Contributions

X. W., H. W., and S. T. designed the study. X. W. carried out the literature search, collected the data and carried out the data analysis. X. W. wrote the manuscript with H. W. and S. T. reviewing and editing. All authors contributed ideas and comments, revised the manuscript, and approved the final version.

## Reference

[1]. COVID-19 dashboard by the center for systems science and engineering at johns hopkins university. Online, 2020 (accessed December28, 2020). https://coronavirus.jhu.edu/map.html.

[2]. Corum J, Grady D, Wee SL, Zimmer C. Coronavirus Vaccine Tracker. Online, 2020 (accessed December28, 2020). https://flowingdata.com/2020/06/10/vaccine-tracker/

[3]. Tang B, Wang X, Li Q, et al. Estimation of the transmission risk of the 2019-nCoV and its implication for public health interventions. J Clin Med 2020; 9:462.

[4]. Tang B, Bragazzi N, Li Q, et al. An updated estimation of the risk of transmission of the novel coronavirus (2019-nCov). Infect Dis Mod 2020; 5:248–255.

[5]. Nishiura H, Kobayashi T, Yang Y, et al. The Rate of Under ascertainment of Novel Coronavirus (2019-nCoV) Infection: Estimation Using Japanese Passengers Data on Evacuation Flights. J Clin Med 2020; 9:E419.

[6]. Zhao S, Musa SS, Lin Q, et al. Estimating the Unreported Number of Novel Coronavirus 2019-nCoV Cases in China in the First Half of January 2020: A Data-Driven Modelling Analysis of the Early Outbreak. J Clin Med 2020; 9:E388

[7]. Kucharski A, Klepac P, Conlan A, et al. Effectiveness of isolation, testing, contact tracing, and physical distancing on reducing transmission of SARS-CoV-2 in different settings: a mathematical modelling study. Lancet Infect Dis 2020; 20(10):1151–1160.

[8]. Karatayeva VA, Anand M, Bauch CT. Local lockdowns outperform global lockdown on the far side of the COVID-19 epidemic curve. PNAS 2020; 117(39):24575–24580.

[9]. Flaxman S, Mishra S, Gandy A, et al. Estimating the effects of non-pharmaceutical interventions on COVID-19 in Europe. Nature 2020; 584:257–261.

[10]. Tang B, Xia F, Tang S, et al. The effectiveness of quarantine and isolation determine the trend of the COVID-19 epidemics in the final phase of the current outbreak in China. Int J Infect Dis 2020; 95:288–293.

[11]. Wang X, Li Q, Sun X, et al. Effects of medical resource capacities and intensities of public mitigation measures on outcomes of COVID-19 outbreaks. medRxiv 2020; doi: https://doi.org/10.1101/2020.04.17.20070318.

[12]. Abbas K, Procter SR, Zandvoort KV, et al. Routine childhood immunisation during the COVID-19 pandemic in Africa: a benefit–risk analysis of health benefits versus excess risk of SARS-CoV-2 infection. Lancet Glob Health 2020; 8(10): e1264–e1272.

[13]. Shen M, Zu J, Fairley CK, et al. Projected COVID-19 epidemic in the United States in the context of the effectiveness of a potential vaccine and implications for social distancing and face mask use. medRxiv 2020; doi: https://doi.org/10.1101/2020.10.28.20221234.

[14]. Bubar KM, Kissler SM, Lipsitch M, et al. Model-informed COVID-19 vaccine prioritization strategies by age and serostatus. medRxiv 2020; doi: https://doi.org/10.1101/2020.09.08.20190629.

[15]. Tang B, Liu P, Yang J, et al. The challenges of the coming mass vaccination and exit strategy in prevention and control of COVID-19, a modelling study. medRxiv 2020; doi: https://doi.org/10.1101/2020.12.18.20248478.

[16]. Matrajt L, Eaton J, Leung T, et al. Vaccine optimization for COVID-19, who to vaccinate first? medRxiv 2020; doi: https://doi.org/10.1101/2020.08.14.20175257.

[17]. Goldstein E, Lipsitch M, Cevik M. On the effect of age on the transmission of SARS-CoV-2 in households, schools and the community. J Infect Dis 2020; 3:3.

[18]. Davies N, Klepac P, Liu Y, et al. Age dependent effects in the transmission and control of COVID-19 epidemics. Nat Med 2020; 26:1205–1211.

[19]. Zhang J, Litvinova M, Liang Y, et al. Changes in contact patterns shape the dynamics of the COVID-19 outbreak in China. Science 2020; 368(6498):1481–1486.

[20]. Keeling M, Tildesley MJ, Atkins BD, et al. The impact of school reopening on the spread ofCOVID-19 in England. medRxiv 2020; doi: https://doi.org/10.1101/2020.06.04.20121434.

[21]. Population Pyramids of the World from 1950 to 2010; 2019. http://www.PopulationPyramid.net.

[22]. Prem K, Cook AR, Jit M. Projecting social contact matrices in 152 countries using contact surveys and demographic data. PLoS Comput Biol 2017; 13(9):e1005697.

[23]. Driessche P, Watmough J. Reproduction numbers and sub-threshold endemic equilibria for compartmental models of disease transmission. Math Biosci 2002; 180:29–48.

[24]. Prem K, Zandvoort K, Klepac P, et al. Projecting contact matrices in 177 geographical regions: an update and comparison with empirical data for the COVID-19 era. medRxiv 2020; doi: https://www.medrxiv.org/content/10.1101/2020.07.22.20159772v2,2020.

[25]. Levin AT, Hanage WP, Owusu-Boaitey N, et al. Assessing the age specificity of infection fatality rates for COVID-19: systematic review, meta-analysis, and public policy implications. Eur J Epidemiol 2020; 35:1123–1138.

[26]. Dooling K, Marin M, Megan W, et al. The advisory committee on immunization practices’ updated interim recommendation for allocation of COVID-19 vaccine — United States, December 2020. MMWR 2020; 51:52. Error! Hyperlink reference not valid.www.cdc.gov/mmwr

