## Supplemental files for "Assessing Age-Specific Vaccination Strategies and Post-Vaccination Reopening Policies for COVID-19 Control Using SEIR Modeling Approach"

**Supplementary Materials**

### The basic reproduction number ($R_0$):

The basic reproduction number is calculated as follows[23]:

The four variables of S, E, A and I are not affected by H, R and V. Therefore, we only consider the four compartments of S, E, A,I, and the infected compartments are E, A and I. So,

$$\mathscr{F}=\begin{pmatrix} p_1^I S_1 \sum_{j=1}^{n} c_{1j} I_j / N + p_1^A S_1 \sum_{j=1}^{n} c_{1j} A_j / N \\ \cdots \\ p_n^I S_n \sum_{j=1}^{n} c_{nj} I_j / N + p_n^A S_n \sum_{j=1}^{n} c_{nj} A_j / N \\ 0 \\ \cdots \\ 0 \end{pmatrix}_{3n\times 1} \quad \mathscr{V}=\begin{pmatrix} \sigma E_1 \\ \cdots \\ \sigma E_n \\ -\sigma(1-\rho_1)E_1+\gamma_A A_1 \\ \cdots \\ -\sigma(1-\rho_n)E_n+\gamma_A A_n \\ -\sigma\rho_1 E_1+\delta I_1 \\ \cdots \\ -\sigma\rho_n E_n+\delta I_n \end{pmatrix}_{3n\times 1}$$

The derivation of the above matricesare obtained

$$F=\begin{pmatrix} O & F_1 & F_2 \\ O & O & O \\ O & O & O \end{pmatrix}, \quad V=\begin{pmatrix} V_1 & O & O \\ V_2 & V_3 & O \\ V_4 & O & V_5 \end{pmatrix}$$

where $$O=\begin{pmatrix} 0 & \cdots & 0 \\ \cdots & \cdots & \cdots \\ 0 & \cdots & 0 \end{pmatrix}, \quad F_1=\frac{1}{N}\begin{pmatrix} p_1^A S_{10} c_{11} & p_1^A S_{10} c_{12} & \cdots & p_1^A S_{10} c_{1n} \\ p_2^A S_{20} c_{21} & p_2^A S_{20} c_{22} & \cdots & p_2^A S_{20} c_{2n} \\ \cdots & \cdots & \cdots & \cdots \\ p_n^A S_{n0} c_{n1} & p_n^A S_{n0} c_{n2} & \cdots & p_n^A S_{n0} c_{nn} \end{pmatrix}$$

$$F_1=\frac{1}{N}\begin{pmatrix} p_1^I S_{10} c_{11} & p_1^I S_{10} c_{12} & \cdots & p_1^I S_{10} c_{1n} \\ p_2^I S_{20} c_{21} & p_2^I S_{20} c_{22} & \cdots & p_2^I S_{20} c_{2n} \\ \cdots & \cdots & \cdots & \cdots \\ p_n^I S_{n0} c_{n1} & p_n^I S_{n0} c_{n2} & \cdots & p_n^I S_{n0} c_{nn} \end{pmatrix}$$

$$V_1=\sigma\begin{pmatrix} 1 & 0 & & 0 \\ 0 & 1 & & 0 \\ & & \cdots & \\ 0 & 0 & & 1 \end{pmatrix}, \quad V_2=-\sigma\begin{pmatrix} 1-\rho_1 & 0 & & 0 \\ 0 & 1-\rho_2 & & 0 \\ & & \cdots & \\ 0 & 0 & & 1-\rho_n \end{pmatrix}, \quad V_3=\gamma_A\begin{pmatrix} 1 & 0 & & 0 \\ 0 & 1 & & 0 \\ & & \cdots & \\ 0 & 0 & & 1 \end{pmatrix}, \quad V_4=-\sigma\begin{pmatrix} \rho_1 & 0 & & 0 \\ 0 & \rho_2 & & 0 \\ & & \cdots & \\ 0 & 0 & & \rho_n \end{pmatrix}$$

$$V_5 = \delta \begin{pmatrix} 1 & 0 & & 0 \\ 0 & 1 & & 0 \\ & & \cdots & \\ 0 & 0 & & 1 \end{pmatrix}.$$

Calculate the inverse of the matrix V, we get $V^{-1} = \begin{pmatrix} V_1^{-1} & O & O \\ -V_3^{-1}V_2V_1^{-1} & V_3^{-1} & O \\ -V_5^{-1}V_4V_1^{-1} & O & V_5^{-1} \end{pmatrix}$.

Therefore, $FV^{-1} = \begin{pmatrix} \Lambda & F_1V_3^{-1} & F_2V_5^{-1} \\ O & O & O \\ O & O & O \end{pmatrix}$,

$$\begin{aligned} \Lambda &= -F_1V_3^{-1}V_2V_1^{-1} - F_2V_5^{-1}V_4V_1^{-1} \\ &= \frac{1}{N\gamma_A} \begin{pmatrix} (1-\rho_1)p_1^A S_{10}c_{11} & (1-\rho_1)p_1^A S_{10}c_{12} & \cdots & (1-\rho_1)p_1^A S_{10}c_{1n} \\ (1-\rho_2)p_2^A S_{20}c_{21} & (1-\rho_2)p_2^A S_{20}c_{22} & \cdots & (1-\rho_2)p_2^A S_{20}c_{2n} \\ \cdots & \cdots & \cdots & \cdots \\ (1-\rho_n)p_n^A S_{n0}c_{n1} & (1-\rho_n)p_n^A S_{n0}c_{n2} & \cdots & (1-\rho_n)p_n^A S_{n0}c_{nn} \end{pmatrix} \\ &+ \frac{1}{N\delta} \begin{pmatrix} \rho_1 p_1^I S_{10}c_{11} & \rho_1 p_1^I S_{10}c_{12} & \cdots & \rho_1 p_1^I S_{10}c_{1n} \\ \rho_2 p_2^I S_{20}c_{21} & \rho_2 p_2^I S_{20}c_{22} & \cdots & \rho_2 p_2^I S_{20}c_{2n} \\ \cdots & \cdots & \cdots & \cdots \\ \rho_n p_n^I S_{n0}c_{n1} & \rho_2 p_n^I S_{n0}c_{n2} & \cdots & \rho_2 p_n^I S_{n0}c_{nn} \end{pmatrix} \end{aligned}$$

The basic reproduction number is the principal eigenvalue of the matrix $\Lambda$.

**Functions of contact rate and detection rate**

In order to accurately describe the variation of control strategies in this model, we assume that the contact rate $c_{ij}(t)$ is decreasing(or increasing) as the increasing (or decreasing) intensity of the control strategy with respect to time *t*. In this study, we focus on three countries, China, India and Italy. The function of $c_{ij}(t)$ for China and India is given by

$$c_{ij}(t) = \begin{cases} c_{ij}^0 & t \le t_c \\ (c_{ij}^0 - c_{ij}^f)e^{-r_c(t-t_c)} + c_{ij}^f & t > t_c \end{cases} \quad (2)$$

where $c_{ij}^0$ denotes the baseline contact rate at the initial time and $c_{ij}^f = q_c c_{ij}^0$ denotes the minimum contact rate under the contact control measures before $t_c$,where $0 \le q_c \le 1$ quantifies the intensity of contact control measures with $q_c$=0indicating the strongest contact control measures to make the final contact rate as 0, and $q_c$=1 indicating “no any effect” of the contact control measures at all. Parameter $r_c$ denotes the exponential decreasing rate of the contact rate after the contact control measures are implemented.

In Italy, the emergency control was established after the first case reported, but shops, theatres and cinemas gradually returned to open in June. In October and November, a number of decrees were issued on strengthening control measures(https://www.acaps.org/covid-19-government-measures-dataset). Thus, the trajectory of the control measures in Italy was initially strong, then relaxed and strong again. Accordingly, we model the function of $c_{ij}(t)$ for Italy as follows,

$$c_{ij}(t)=\begin{cases}(c_{ij}^{0}-c_{ij}^{f_1})e^{-r_{c_1}(t)}+c_{ij}^{f_1} & t\le t_{c_1}\\ (c_{ij}^{f_1}-c_{ij}^{f_2})e^{-r_{c_2}(t-t_{c_1})}+c_{ij}^{f_2} & t_{c_1}<t\le t_{c_2}\\ (c_{ij}^{f_2}-c_{ij}^{f_3})e^{-r_{c_3}(t-t_{c_2})}+c_{ij}^{f_3} & t>t_{c_2}\end{cases} \qquad (3)$$

where $c_{ij}^{f_1}$ ($c_{ij}^{f_1}=q_{c_1}c_{ij}^{0}$), $c_{ij}^{f_2}$ ($c_{ij}^{f_2}=q_{c_2}c_{ij}^{f_1}$)and $c_{ij}^{f_3}$ ($c_{ij}^{f_3}=q_{c_3}c_{ij}^{f_2}$) are the minimum or the maximum contact rate under control strategies or due to relaxation of control. $r_{c_1}$, $r_{c_2}$ and $r_{c_3}$ denote how an exponential increase or decrease in the contact rate is affected by strengthening control or relaxation of control, $t_{c_1}$ and $t_{c_2}$ are the switching time of control strength.

We also set the transition rate $\delta(t)$ as an increasing function with respect to time t, with the following form:

$$\delta(t)=\begin{cases}\delta_0 & t\le t_c\\ (\delta_0-\delta_f)e^{-r_\delta(t-t_c)}+\delta_f & t>t_c\end{cases} \qquad (4)$$

where $\delta_0$ is the initial rate of confirmation(detection), $\delta_f$ is the fastest confirmation rate ($\delta_f=q_\delta\delta_0, q_\delta>1$), and $r_\delta$ is the exponentially increasing rate. The critical time $t_c$ for China is January 23th, 2020when Wuhan city and all parts of the country continued to take stringent control measures. The onset time of the epidemic in Italy and India was later than January 23, 2020, so we set $t_c$ for India and Italy to be 0.

**Parameter estimation**

Other parameters related to control measures such as the variation of contact rate and detection rate, and some initial conditions such as initial values of the exposed individuals and infected individuals are estimated by fitting the model to the daily reported cases(Fig.1 and Table 1)by the nonlinear least squares (NLS) estimation method. We assume that the measurement error of the data(the daily confirmed cases $Y(t)$) follows a distribution with mean 0 and variance $\sigma^2$. The NLS objective function is

$$L(\cdot)=\sum_{t=1}^{T}(Y(t)-\sum_{i=1}^{16}\delta I_i)^2 \qquad (6)$$

where $T$ is the length of the data used for model fitting. The interior-point method was used to optimize the loss function (6), and implemented with MATLAB. Here, since there are few exposed and infected persons at the beginning of the epidemic, we assume that the number of exposed persons and the number of infected persons(asymptomatic and symptomatic) in different age groups are equal.

**Model fitting and parameter estimation results**

Fig.1 shows the data of daily confirmed new cases (circle) and the results of model fitting (solid curve) for the three countries, China, India and Italy. We can see that the fitted models capture the trends of the observed data very well. In particular, a small wave and a big peak of the epidemic in Italy were captured on March 21 and November 13 respectively(Figure 1b), and a single wave of the epidemic in China and India was captured on February 12 and September 16 by the model (Figure 1a and 1c). The estimated model parameters as well as the derived parameters from literature are shown in Table 1.

### Figures

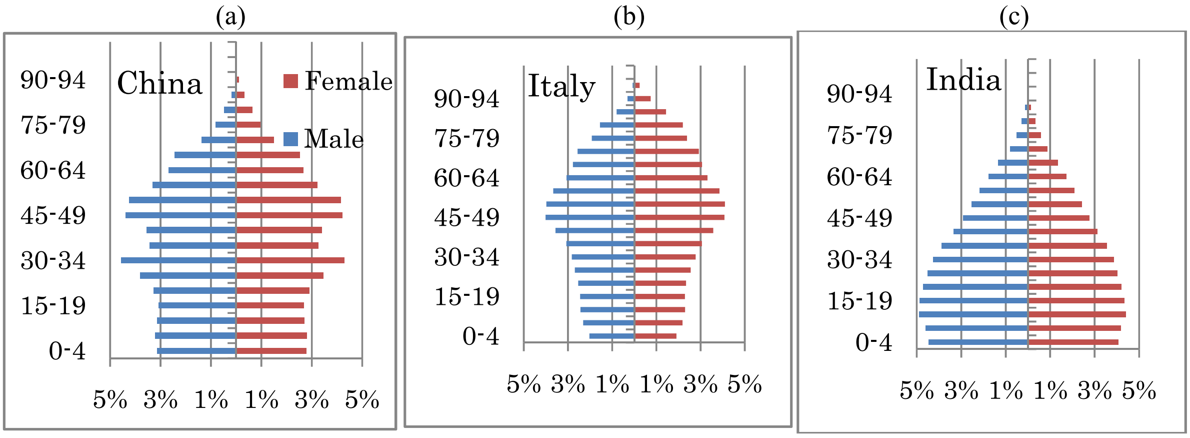


Fig.S1: The population age distribution for the three countries (China, Italy and India).

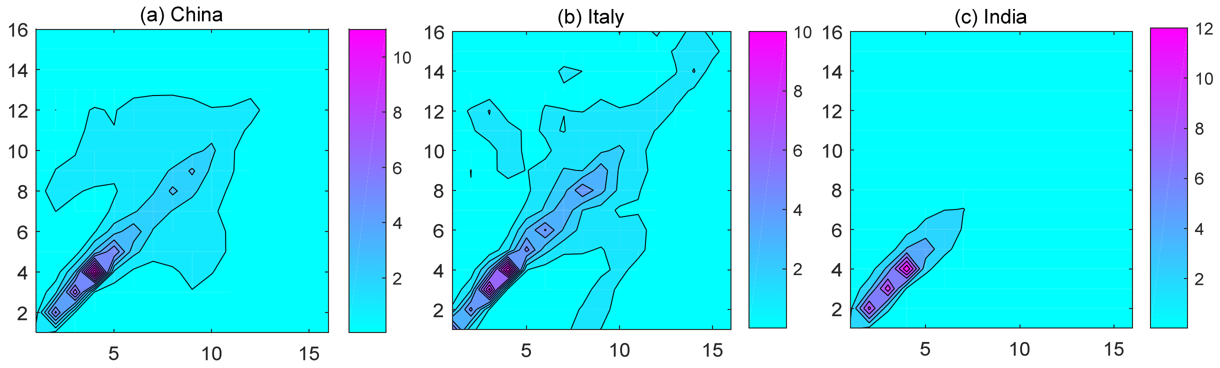


Fig.S2: Thecontact pattern of the three countries (China, Italy and India) for different age groups.

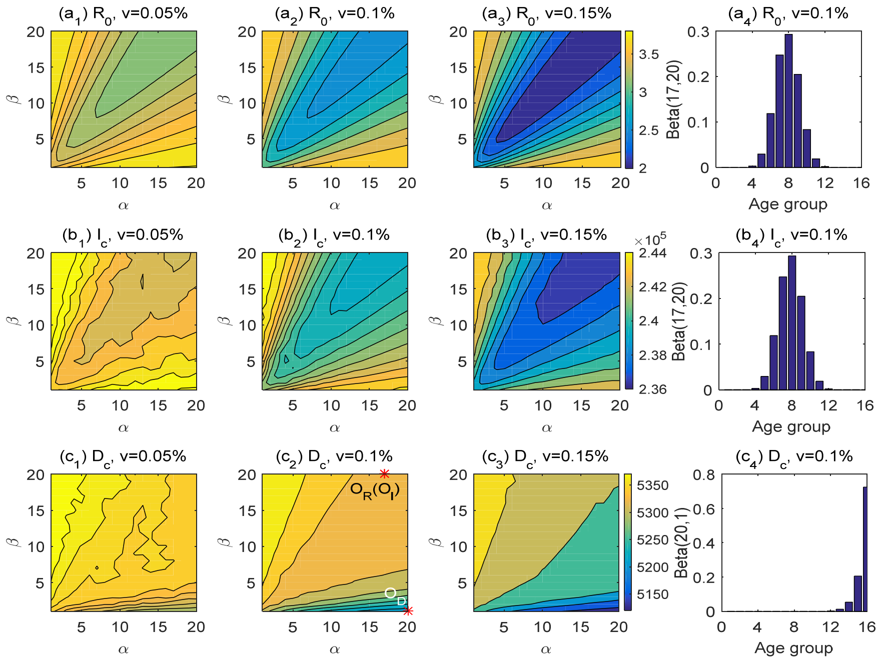


Fig.S3: The contour plot of the three endpoints: the basic reproduction number ($R_0$, 1st row), the cumulative number of infections ($I_c$, 2nd row) and the cumulative number of deaths ($D_c$, 3rd row) for Italy. The optimal age-specific vaccination distributions for these three endpoints are shown in ($a_4$), ($b_4$) and ($c_4$) respectively when $v = 0.1\%$. $O_R$, $O_I$, and $O_D$ are the optimal points obtained by minimizing the three endpoints ($R_0$, $I_c$, $D_c$) respectively.

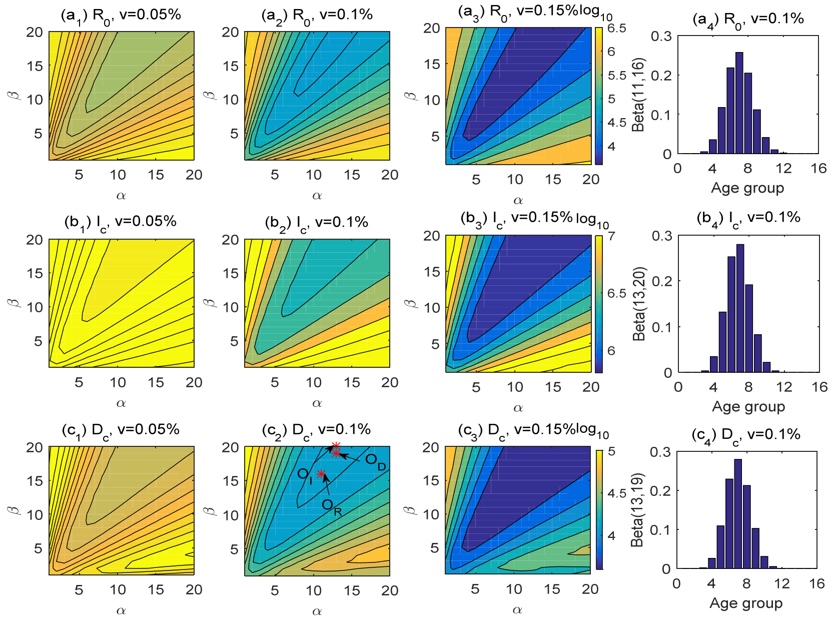


Fig.S4: The contour plot of the three endpoints: the basic reproduction number ($R_0$, 1st row), the cumulative number of infections ($I_c$, 2nd row) and the cumulative number of deaths ($D_c$, 3rd row) for China. The optimal age-specific vaccination distributions for these three endpoints are shown in ($a_4$), ($b_4$) and ($c_4$)respectively when $v = 0.1\%$. $O_R$, $O_I$,and $O_D$ are the optimal points obtained by minimizing the three endpoints ($R_0$, $I_c$, $D_c$) respectively.

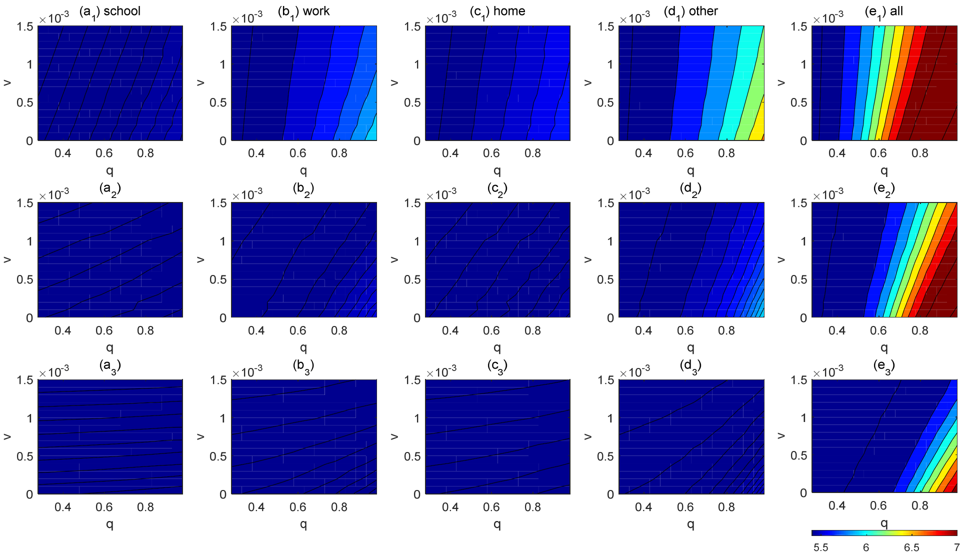


Fig.S5: Simulation results for the number of infections (in log10 scale) for Italy during the time period [T+1,T+180] for five different scenarios of contact control release (a-e) for different values of $q$ and $v$ under different releasing times,T+1 (1st row), T+30(2nd row), and T+60 (3rd row). The optimal age-specific vaccination strategy with the initiation time of vaccination as $T+1$ was assumed for all the simulation scenarios.

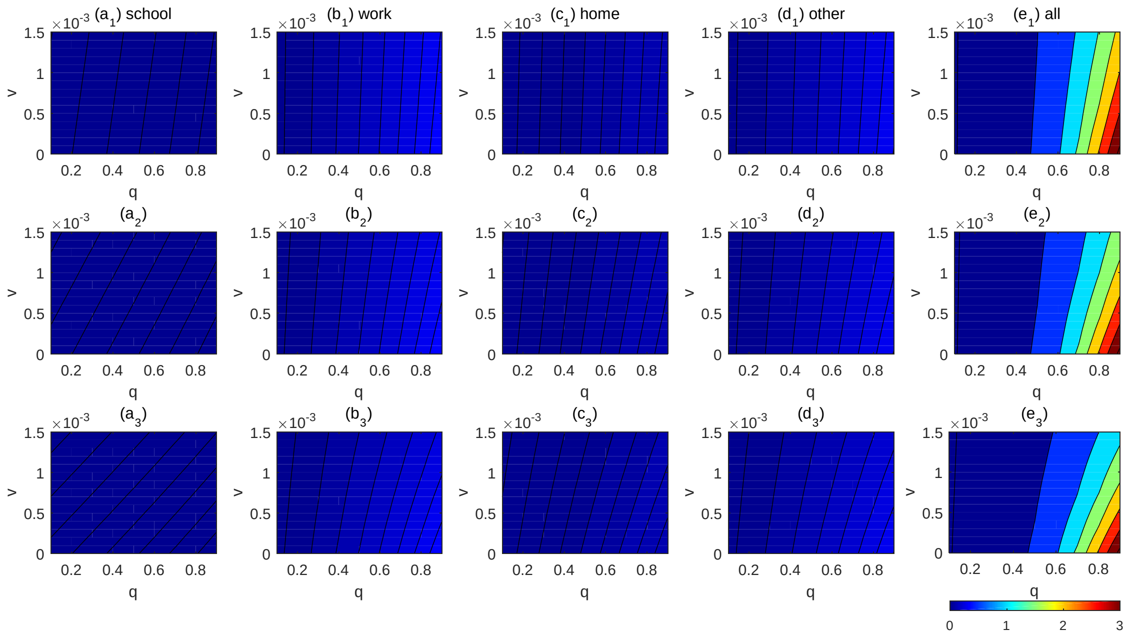


Fig.S6: Contour plot of the number of infections (log10 scale) as a function of the release ratio of contact rate $q$ ($c_{ij}(t) = qc_{ij}^{0}, t > T$) and the daily vaccination rate v for China within 6 months after releasing under different initial releasing time: T+1 (the first line), T+30(the second line), T+60 (the third line).We assumedone infected individual is imported and the detection rate $\delta = 0.4$.

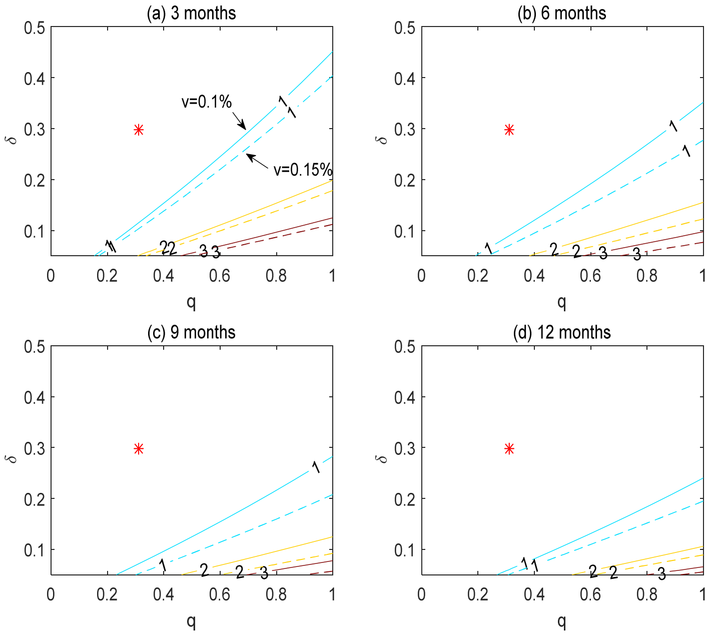


Fig.S7: Contour plot of the basic reproduction number ($R_0$)as a function of the contact control release rate (q) and the detection rate (δ) with different daily vaccination rates v=0.1% (the solid line) and v=0.15%(the dash line)under the optimal age-specific vaccination strategy in Italy. The vaccination periods are set to be 3 months(a), 6 months(b), 9 months (c) and 12 months (d). The red dot is the current value of these two parameters.

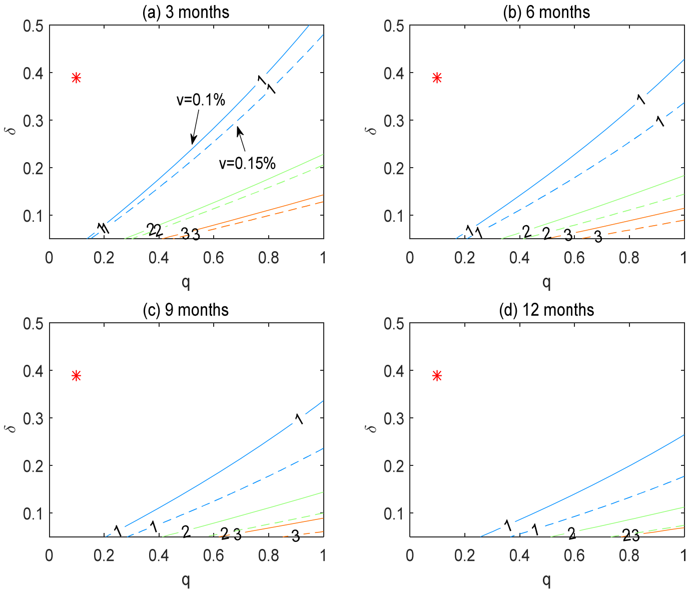


Fig.S8: Contour plot of the basic reproduction number ($R_0$)as a function of the contact control release rate (q) and the detection rate (δ) with different daily vaccination rates v=0.1% (the solid line) and v=0.15%(the dash line)under the optimal age-specific vaccination strategy in China. The vaccination periods are set to be 3 months(a), 6 months(b), 9 months (c) and 12 months (d). The red dot is the current value of these two parameters.

**Tables:**

Table S1: Optimal age-specific vaccination distributions for different daily vaccination rates and different endpoints for three countries (China, Italy and India).

| China | R | Cumulative infection | Cumulative death |
|---|---|---|---|
| 0.05% | Beta(13,20) | Beta(13,20) | Beta(14,20) |
| 0.15% | Beta(8,11) | Beta(12,18) | Beta(12,17) |
| Italy | R | Cumulative infection | Cumulative death |
| 0.05% | Beta(17,20) | Beta(17,20) | Beta(16,1) |
| 0.15% | Beta(17,20) | Beta(16,18) | Beta(20,1) |
| India | R | Cumulative infection | Cumulative death |
| 0.05% | Beta(7,20) | Beta(7,20) | Beta(20,1) |
| 0.15% | Beta(5,13) | Beta(8,20) | Beta(20,1) |
